# A surrogate virus neutralization test to quantify antibody-mediated inhibition of SARS-CoV-2 in finger stick dried blood spot samples

**DOI:** 10.1101/2021.02.14.21251709

**Authors:** Amelia Sancilio, Richard D’Aquila, Elizabeth M. McNally, Matt E Velez, Michael G. Ison, Alexis R. Demonbreun, Thomas W. McDade

## Abstract

**Background:** The spike protein of SARS-CoV-2 engages the human angiotensin-converting enzyme 2 (ACE2) receptor to enter host cells, and neutralizing antibodies are effective at blocking this interaction to prevent infection. Widespread application of this important marker of protective immunity is limited by logistical and technical challenges associated with live virus methods and venous blood collection. To address this gap, we validated an immunoassay-based method for quantifying neutralization of the spike-ACE2 interaction in a single drop of capillary whole blood, collected on filter paper as a dried blood spot (DBS) sample.

**Methods:** Samples are eluted overnight and incubated in the presence of spike antigen and ACE2 in a 96-well solid phase plate. Competitive immunoassay with electrochemiluminescent label is used to quantify neutralizing activity. The following measures of assay performance were evaluated: dilution series of confirmed positive and negative samples, agreement with results from matched DBS-serum samples, analysis of results from DBS samples with known COVID-19 status, and precision (intra-assay percent coefficient of variation; %CV) and reliability (inter-assay; %CV).

**Results:** Dilution series produced the expected pattern of dose-response. Agreement between results from serum and DBS samples was high, with concordance correlation = 0.991. Analysis of three control samples across the measurement range indicated acceptable levels of precision and reliability. Median % neutralization was 46.9 for PCR confirmed convalescent COVID-19 samples and 0.1 for negative samples.

**Conclusions:** Large-scale testing is important for quantifying neutralizing antibodies that can provide protection against COVID-19 in order to estimate the level of immunity in the general population. DBS provides a minimally-invasive, low cost alternative to venous blood collection, and this scalable immunoassay-based method for quantifying neutralization of the spike-ACE2 interaction can be used as a surrogate for virus-based assays to expand testing across a wide range of settings and populations.

## INTRODUCTION

Coronavirus disease 2019 (COVID-19) is a potentially deadly infectious disease that is caused by the severe acute respiratory syndrome coronavirus 2 (SARS-CoV-2)^1^. Serological testing is an important part of the effort to slow the spread of SARS-CoV-2, and detects the presence of antibodies in blood samples from exposed individuals ^2^. Antibodies are detectable 3-10 days after infection, and anti-SARS-CoV IgG antibodies persist for at least 5 months after the resolution of symptoms ^3,4^. Neutralizing antibodies (NAbs) are of particular importance because they can bind to viral proteins and inhibit entry into host cells, thereby preventing infection ^5^. For SARS-CoV-2, the receptor binding domain (RBD) of the surface spike protein engages the human angiotensin-converting enzyme 2 (ACE2) receptor to gain entry into host cells, and anti-SARS-CoV-2 NAbs are effective at blocking this interaction and preventing viral entry ^6,7^.

Serological testing has been widely used to identify individuals who have been exposed to SARS-CoV-2. Importantly, due to their functional role in blocking viral entry, the measurement of NAbs provides information on the magnitude of immune response to prior infection and the level of protection against re-infection ^8^. In addition, NAbs can be used to assess conferred immunity through vaccination, as neutralizing antibody response is frequently used as a correlate of protection following vaccination, including vaccination against SARS-CoV-2 ^9,10^.

Conventional methods for measuring neutralizing activity require live virus—either a non-replicating virus with the SARS-CoV-2 spike protein expressed from a viral vector (“pseudotyped virus” or “pseudovirus”) or replication-competent SARS-CoV-2. Either live virus approach limits widespread application as the assays are time-consuming and require specialized laboratory containment facilities. SARS-CoV-2 neutralizing antibodies can also be quantified utilizing a surrogate virus neutralization test (sVNT), which detects NAbs without live virus and can be implemented with immunoassay techniques ^6,11^. The sVNT approach replicates the virus-host interaction by incubating a blood sample with purified versions of viral spike protein and human ACE2 receptor. Neutralizing antibodies prevent the spike protein from binding to ACE2, and the degree of inhibition is quantified in a competitive immunoassay to indicate the level of NAbs in the sample. There is a high level of agreement in results derived from sVNT and live virus methods, and the specificity and sensitivity of sVNT are comparable to conventional neutralization tests ^11,12^.

While sVNT provides a more streamlined alternative to conventional assays for neutralizing antibodies, the use of venous blood limits the screening of large numbers of people in non-clinical settings due to the costs and logistical constraints associated with collecting, processing, and transporting venous blood. There is an alternative in finger stick dried blood spot (DBS) sampling^13^. With DBS, a lancet is used to prick the finger, and up to five drops of blood are placed on filter paper, where it dries. In comparison with venipuncture, DBS sampling is low-cost, reduces biohazard risk during sample collection and transport, and it affords the possibility of self-sampling, with participants collecting their own blood and returning the sample through the mail, without the need for a cold chain or special handling^14^. Due to these advantages, DBS sampling is the foundation of neonatal screening programs in the US, and it is increasingly applied as a minimally-invasive alternative to venipuncture in community- and population-based health research, including several applications in infectious disease epidemiology ^15,16^.

We recently validated a protocol for quantifying anti-RBD SARS-CoV-2 IgG antibodies in DBS samples that is being used in several community-based seroprevalence studies ^17-19^. Here, we describe a method for quantifying neutralization of spike-ACE2 binding in DBS samples.

The protocol is relatively short (<4 hours), can be implemented with widely available laboratory infrastructure, and can be scaled up for high-throughput processing. We describe the assay protocol, as well as results demonstrating performance characteristics and the validity of measuring SARS-CoV-2 neutralizing antibodies in DBS samples.

## MATERIALS AND METHODS

The DBS method is based on modifications to a commercially available protocol (Meso Scale Diagnostics V-PLEX SARS-CoV-2 Panel 2 Kit; K15386U-2), designed for the multiplex capture of SARS-CoV-2 antibodies in serum on the Meso Scale Diagnostics QuickPlex SQ 120 Imager. This method utilizes a 96-well solid phase plate with antigens printed as arrays in discrete spots within each well, an electrochemiluminescent detection system, and a competitive binding protocol. The assay principle is as follows: Diluted sample is incubated in plate wells in the presence of ACE2 conjugated with an electrochemiluminescent label; neutralizing antibodies, if present, compete with conjugated ACE2 for viral antigen binding sites; after separation an electrical impulse generates light in proportion to bound ACE2 at each spot in the array; the signal is negatively proportional to the concentration of neutralizing antibodies in the sample, and % neutralization is calculated as follows: % neutralization = 100 x 1 - (sample signal/negative control signal).

### Serum assay protocol

Serum samples were quantified per manufacturer’s instructions. Briefly, the assay plate included two SARS-CoV-2 antigens coated to the bottom of each well: spike and RBD. The nucleocapsid antigen is also available in the plate, but does not bind ACE2 and is not a target for neutralizing antibodies; it was therefore not used in this protocol. The assay plate was blocked for 30 minutes and washed. Serum samples were diluted (1:40) and 25uL were transferred to each well. The plate was then incubated at room temperature for 60 minutes while shaking at 700rpm, followed by the addition of SULFO-TAG^™^ conjugated ACE2, and continued incubation with shaking for 60 minutes. The plate was washed, 150 μL MSD GOLD^™^ Read Buffer B was added to each well, and the plate was read using the QuickPlex SQ 120 Imager. Mean fluorescence intensity (MFI) values were generated for each sample. Sample diluent was used as the negative control for the calculation of % neutralization.

### DBS assay protocol

Three levels of DBS control material (low, mid, and high neutralization antibodies) were manufactured by diluting neutralizing monoclonal antibody to SARS CoV-2 spike (provided with kit) in assay diluent, mixing with an equal volume of washed red blood cells, then transferring to filter paper (Whatman 903 Protein Saver Card, #10534612) in 60 uL drops using a pipette (Rainin pipet lite LTS L100)^13^. A DBS negative control sample was made with the same procedure, using assay diluent.

To begin the protocol, DBS samples and DBS controls were eluted overnight in a non-binding 96-well plate (Corning 96 Well TC-Treated Microplate; Sigma #CLS3599). One 5.0 mm disc was removed using a pneumatic hole punch (Analytical Sales and Services #327500, Flanders, NJ), 100 uL assay diluent were added to each well, and the plate was covered and incubated overnight at 4°C.

The following day, the elution plate was from removed from the refrigerator and rotated for one hour at 300 rpm. The MULTI-SPOT® 96-well assay plate was blocked for 30 min at room temperature, washed, and 25 uL of each sample were transferred from the elution plate to the assay plate in duplicate. The assay plate was then incubated at room temperature for 60 minutes while shaking at 700rpm, followed by the addition of 25 uL of SULFO-TAG^™^ conjugated ACE2, and continued incubation with shaking for 60 minutes. The plate was washed, 150 μL MSD GOLD^™^ Read Buffer B was added to each well, and the plate was read using a MESO® QuickPlex SQ 120MM Imager. Mean MFI values were generated for each sample, and the DBS negative control was used to calculate % neutralization for each sample.

### Evaluation of assay performance

The following aspects of DBS assay performance were evaluated: dilution series of confirmed positive and negative samples, agreement with results from serum samples, analysis of results from DBS samples with known COVID-19 status, and precision and reliability.. Assay precision, or intra-assay variability, was calculated as the percent coefficient of variation (CV; SD/mean x 100) for 10 replicates each of DBS control samples with high, medium, and low levels of neutralizing antibodies. Reliability, or inter-assay variability, was assessed by running the three DBS control samples across ten separate assay plates.

Agreement between results from DBS and serum samples was evaluated in a set of matched samples collected from n=20 convalescent individuals with PCR positive confirmed cases of COVID-19, and n=10 negative samples collected in early 2019, before the pandemic. Immediately following collection of venous blood, five drops of whole blood (60 uL each) were transferred by pipet to filter paper (Whatman #903, GE Healthcare, Pascataway, NJ), allowed to dry overnight, and then stacked and stored at -30°C. DBS samples with known SARS-CoV-2 seropositivity status were drawn from an ongoing community-based seroprevalence study and collected between June 24 and November 11, 2020, with serostatus determined using a SARS-CoV-2 anti-RBD IgG immunoassay validated for use with DBS ^17,18^. These samples were categorized as seronegative (n=38), seropositive with PCR confirmed COVID-19 (n=51), and seropositive with no PCR test (n=32). An additional set of negative DBS samples, collect in early 2019, were also assayed (n=27). All samples were de-identified and all research activities were implemented under protocols approved by the institutional review board at Northwestern University (#STU00212371, #STU00212457 and #STU00212472).

All samples were analyzed in duplicate using the DBS and serum protocols, as described above. Passing-Bablok regression models and difference plots were used to evaluate patterns of agreement in results ^24-27^. Statistical analyses were implemented with Stata/SE (version 15.1 for Windows, StataCorp, College Station, TX). Results are presented for neutralization of the spike-ACE2 interaction only. Results for neutralization of RBD-ACE2 interaction were highly correlated with spike (Spearman R = 0.90) and were not considered further as part of this validation.

## RESULTS

Analysis of DBS samples from convalescent, PCR confirmed cases of COVID-19 demonstrated that the SARS-CoV-2 Spike-ACE2 interaction can be neutralized in a dose-dependent manner (Figure 1). There was no evidence of neutralization in pre-pandemic negative DBS samples. Results from a set of matched DBS and serum samples, including confirmed positive and negative cases, indicated a high level of agreement in % neutralization across the two sample types (concordance correlation = 0.991) (Figure 2). A Bland-Altman difference plot indicated that the pattern of association was linear and consistent across the measurement range, with relatively tight 95% limits of agreement (bias = 0.63 % inhibition; lower limit = -8.4; upper limit = 9.6) (Figure 3).

**Figure 1.**
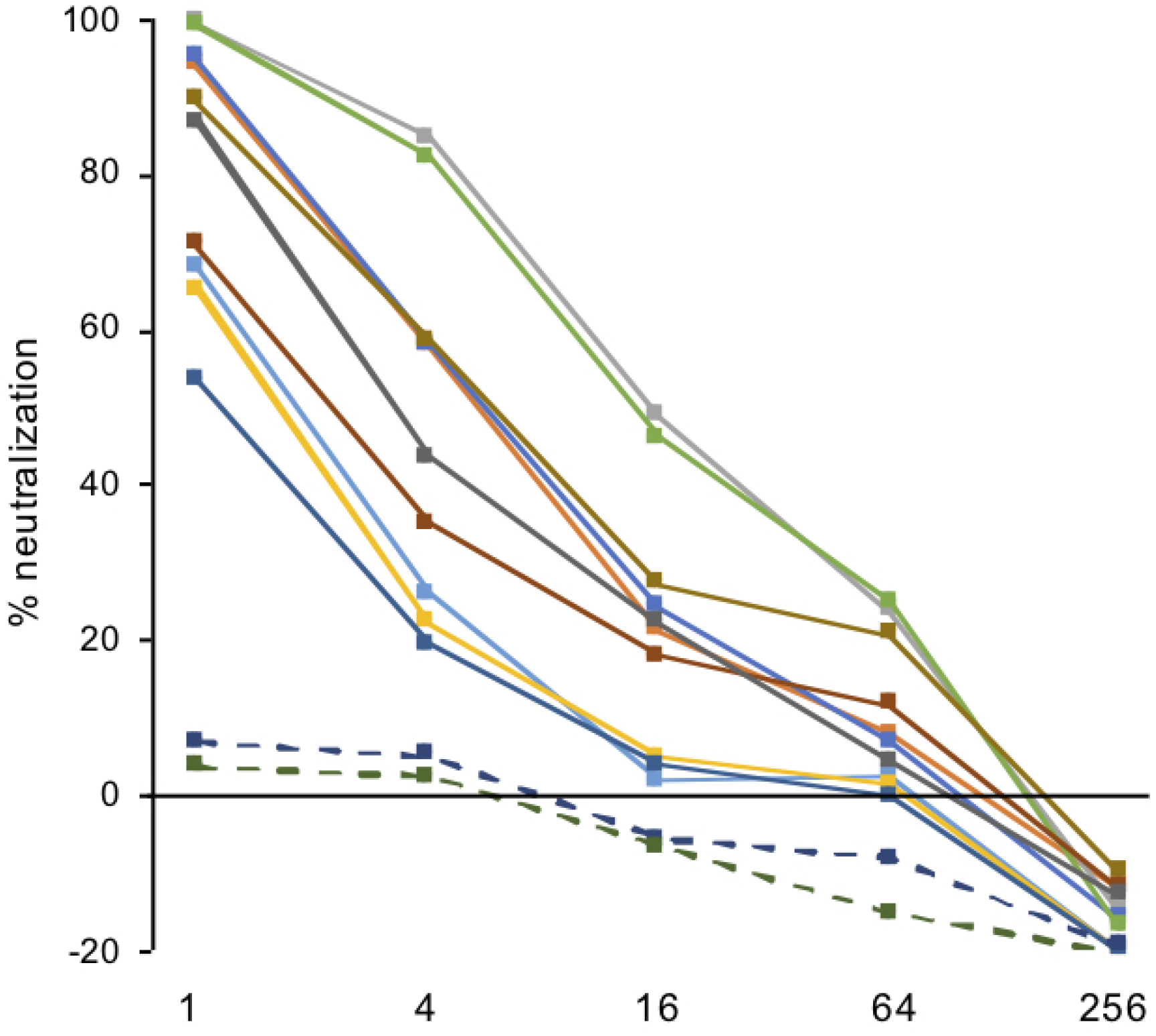
Neutralization of SARS-CoV-2 Spike-ACE2 interaction in DBS samples from convalescent COVID-19 cases and pre-pandemic negative samples. Reduction in neutralization with dilution of DBS samples (1:4, 1:16, 1:64, 1:256) after initial elution of one 5 mm punch in 100 uL assay buffer. Negative samples are indicated with dashed lines.

**Figure 2.**
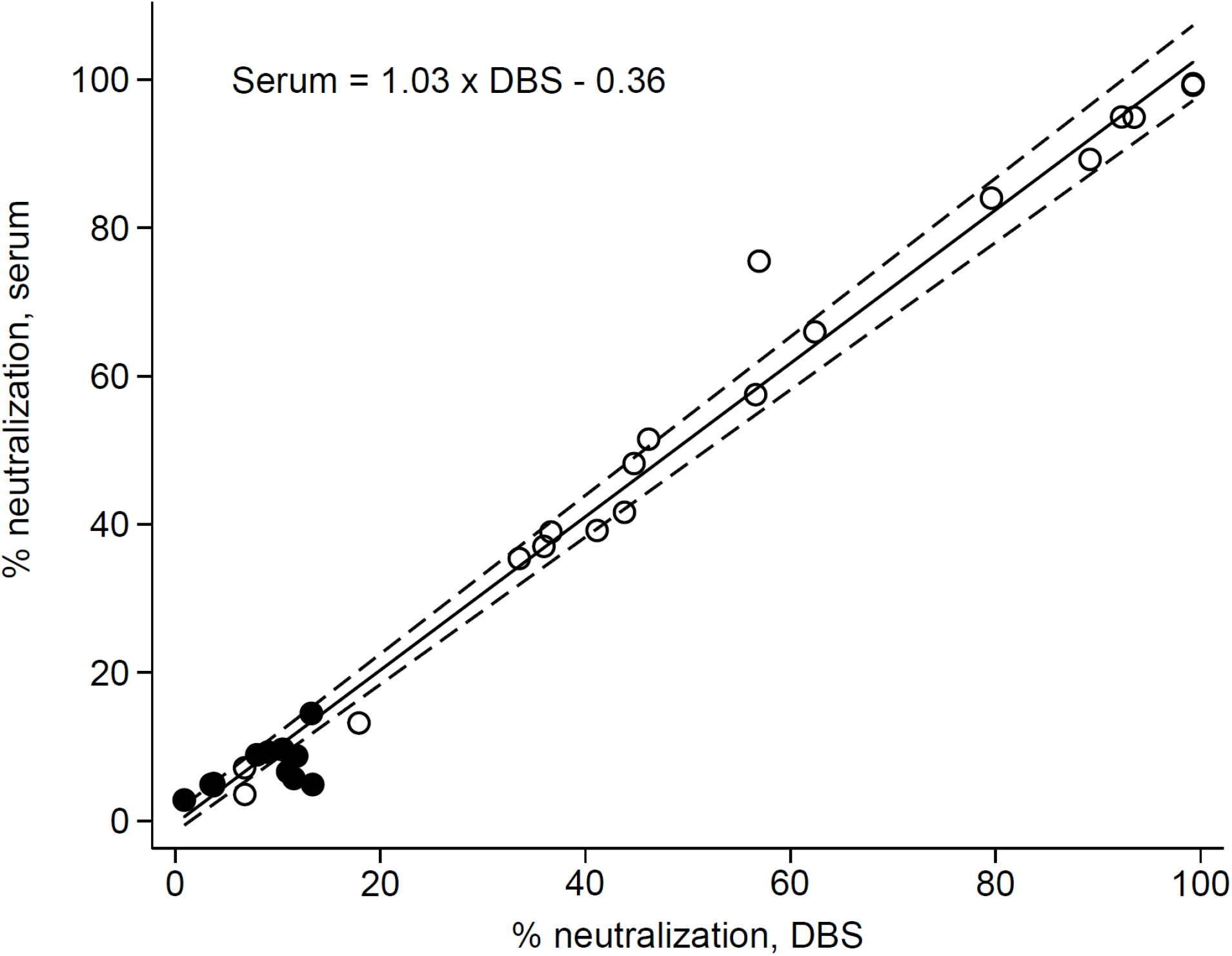
Agreement in neutralization results for matched serum and DBS samples. Scatterplot and Passing-Bablok regression line (95% CI) demonstrating agreement in % neutralization of SARS-CoV-2 Spike-ACE2 interaction in DBS and serum samples. PCR positive samples are indicated with open circles; negative samples are indicated with black circles. The concordance correlation of absolute agreement = 0.991.

**Figure 3.**
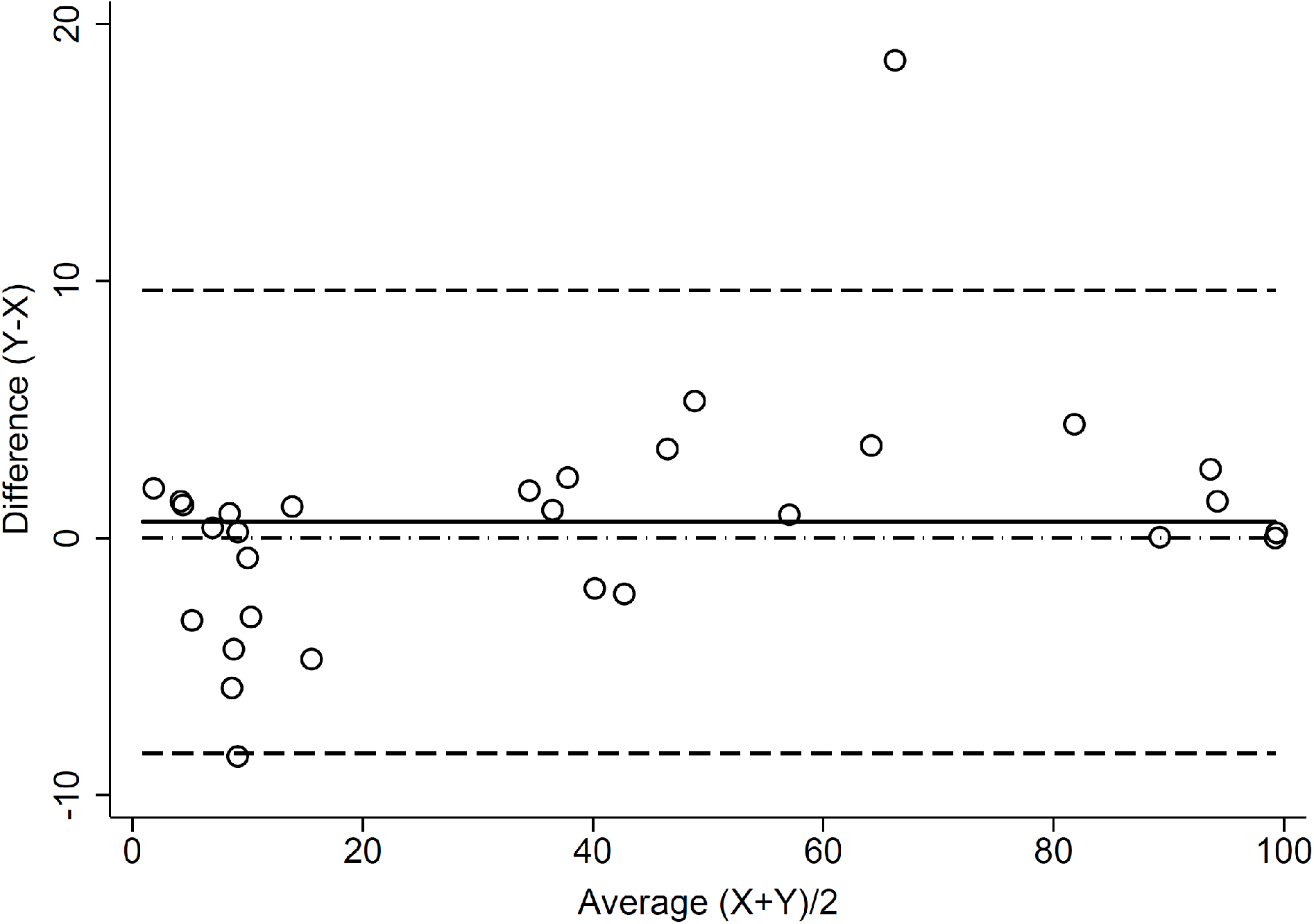
Agreement in neutralization results for matched serum and DBS samples.Bland-Altman difference plot with 95% limits of agreement for results from serum and DBS samples for % neutralization of SARS-CoV-2 spike-ACE2 interaction (X=DBS; Y=serum). Mean bias = 0.63 (95% CI: -1.09, 2.34); lower limit of agreement = -8.38 (95% CI: -11.34, -5.41); upper limit of agreement = 9.64 (95% CI: 6.66, 12.61); concordance correlation of absolute agreement = 0.991.

Repeat analysis of three Nab-positive DBS control samples indicated a high level of precision and reliability for the assay, particularly at higher levels of neutralization of the spike-ACE2 interaction. Levels of intra-assay variability for high, mid, and low Nab positive DBS control samples were 0.2, 3.2, and 14.9 %CV, respectively. Inter-assay variability was 0.7, 5.7, and 14.1 %CV, respectively.

Analysis of DBS samples with known COVID-19 serostatus produced the expected pattern of results, with the highest levels of neutralization in samples from convalescent, PCR confirmed cases (Figure 4). Median neutralization in this group was 46.9%, but values spanned the entire measurement range. All negative samples (seronegative and pre-pandemic) showed low levels of neutralization, with % neutralization <14%. Median % neutralization was 0.1% for negative samples, and mean neutralization was 0.0% (SD=6.5). Using 3SD above the mean as a criterion, the cut-off value for differentiating negative from positive samples is 19.5%.

**Figure 4.**
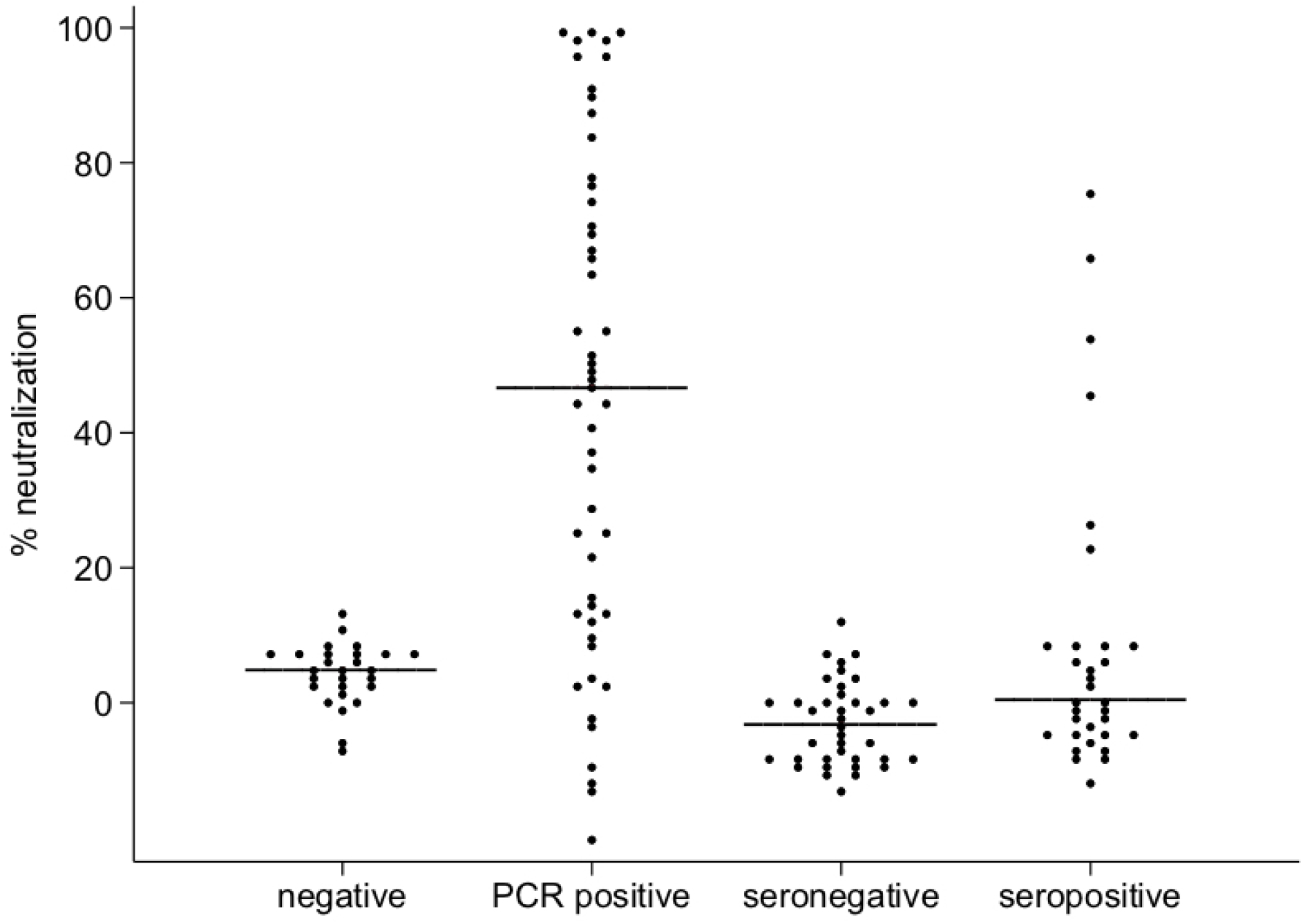
Neutralization of SARS-CoV-2 Spike-ACE2 interaction in DBS samples. Negative samples were collected in 2019; PCR positive samples are from convalescent COVID-19 cases. Seronegative and seropositive samples were community-acquired between June 24 and November 11, 2020, with serostatus determined based on the presence of SARS-CoV-2 anti-RBD IgG antibodies. The black line represents median neutralization for each group.

Neutralizing activity that blocks spike-ACE2 interaction in this assay above this threshold was detected in only 18.8% of samples collected from individuals who tested seropositive for SARS-CoV-2 anti-RBD IgG but who did not have symptoms that led to a clinical diagnosis of COVID-19. Among confirmed PCR positive cases, 68.6% were above the cut-off.

## DISCUSSION

Serological testing is critical for tracking the spread of SARS-CoV-2, and for identifying the factors that influence transmission, immunity, and vulnerability to severe infection. High rates of asymptomatic and mild infection underscore the need for approaches that are amenable to large-scale application. Testing for neutralizing antibodies may be particularly important at this stage in the pandemic, as it can inform our understanding of immunity among vaccinated individuals and those previously exposed to SARS-CoV-2. We have validated a method for quantifying neutralization of the SARS-CoV-2 spike-ACE2 interaction in a single drop of blood, collected on filter paper following a simple finger stick, in order to facilitate remote sampling and large-scale testing of neutralizing antibodies.

Our results demonstrate the feasibility of using DBS samples for serological testing of SARS-CoV-2 neutralizing antibodies with a competitive immunoassay that quantifies inhibition of the spike-ACE2 interaction. Neutralization is high in samples from PCR confirmed convalescent cases of COVID-19, and low in samples that are known to be negative for SARS-CoV-2. We document dose-dependent reduction in neutralization in dilutions of samples with high initial neutralization, as well as close correspondence in results from DBS in comparison with matched serum samples. The method also produces results that are precise and reliable, which is particularly important for studies with large numbers of samples assayed across multiple plates. Furthermore, the method can be implemented in ubiquitous BSL2 laboratories, without the specialized facilities and biohazard risks associated with live virus methods.

Finger stick DBS sampling is relatively low-cost and minimally-invasive, and samples can be self-collected in the home ^14,15^. In addition, antibodies remain stable in DBS for several weeks at room temperature, and can be sent in the mail as nonregulated, exempt materials ^20^. The possibility of remote serological testing, with large numbers of people, may be particularly advantageous when clinical resources are stretched thin and people are being encouraged to stay home. A disadvantages of DBS includes the small volume of blood in comparison with venipuncture. Other potential issues, depending on the application, are variable quality of self-collected samples, and the possibility that samples may be delayed or lost during shipment by mail.

Recent studies have documented neutralization antibody responses to vaccination and natural infection with SARS-CoV-2 ^8,21,22^, and prior research with other viruses suggests that neutralizing antibodies are excellent indicators of protective immunity ^9^. More widespread, community-based testing is important for determining the level of neutralizing antibodies that provide protection against COVID-19, and for estimating the level and duration of immunity in the general population—and subpopulations of particular interest—following multiple waves of community transmission ^23^. Testing for neutralization antibodies can also play a role in evaluating the effectiveness of vaccines, and heterogeneity in vaccine responses across individuals ^8^. Our quantitative DBS-based assay for neutralizing antibodies can help advance research and surveillance in these important areas.

## Data Availability

Data are available upon request from the corresponding author.

